# Polygenic risk scores associate with asthma phenotypes and proteomic analyses implicate IL1R1 in two family-based studies

**DOI:** 10.64898/2026.06.06.26355045

**Authors:** Sanghun Lee, Matthew Moll, Kevin Mendez, Nicole Prince, Jessica Lasky-Su, Sharon M. Lutz, Scott T. Weiss, Christoph Lange, Rachel S. Kelly, Julian Hecker

## Abstract

Despite its high prevalence and the discovery of hundreds of genetic associations, the genetic determinants and heterogeneous manifestations of asthma remain incompletely understood. Incorporating polygenic risk scores (PRS) into asthma research offers a powerful approach to quantify inherited susceptibility, refine risk profiles, and advance mechanistic understanding of disease development. For this study, we leveraged whole-genome sequencing (WGS) data from two family-based cohorts of childhood asthma - the Genetics of Asthma in Costa Rica Study (GACRS) and the Childhood Asthma Management Program (CAMP) - to examine the transmission profiles of externally derived asthma PRS and their associations with clinical phenotypes in children with asthma. To further elucidate molecular mechanisms, we integrated large-scale external genome-wide association study (GWAS) summary statistics and genetic prediction models of protein abundance in a two-step proteome-wide association study (PWAS) of asthma. Our findings provide robust evidence supporting the validity of externally derived asthma PRS (asthma PRS association p-value *p* = 10^−24^ [GACRS and CAMP trios combined] for the Global Biobank Meta-analysis Initiative [GBMI]) and reveal consistent associations with spirometry measures and atopy markers across both studies, as 13 of 21 traits (62%) were significantly associated with the GBMI-PRS in the meta-analysis after multiple-testing correction. Moreover, the results of the integrative proteomic analysis implicate IL-1 signaling in the etiology of asthma, reinforcing the candidacy of IL1R1 antagonists for drug repurposing.

## Introduction

Asthma is a common chronic respiratory disease that affects more than 300 million people worldwide, contributing substantially to global morbidity and healthcare costs^1^. Asthma is characterized by variable airflow obstruction, airway hyperresponsiveness (AHR), airway inflammation, and an age-at-onset-specific genetic architecture^2^, with clinical manifestations ranging from wheezing and cough to severe exacerbations^3^. Early family and twin studies have consistently demonstrated a substantial genetic contribution to asthma^4^. Over the past two decades, genome-wide association studies (GWAS) have identified more than a hundred genetic loci associated with asthma susceptibility, including the well-established *ORMDL3/GSDMB* locus on 17q21^5^. Despite these advances, the identified genetic effects account for only a fraction of asthma heritability, and their translation into clinically meaningful tools has been limited.

Polygenic risk scores (PRS), which aggregate the effects of thousands of common variants into a single measure of genetic liability, have emerged as powerful tools for studying the genetic profiles of complex diseases and their comorbidities, including asthma. Large-scale efforts such as the Trans-National Asthma Genetic Consortium (TAGC) and the Global Biobank Meta-analysis Initiative (GBMI) have generated valuable resources to develop robust PRS for asthma^5,6^. However, most evaluations of PRS to date have been conducted in population-based or case– control settings, and studies using family-based designs are lacking. Family-based designs enable genetic analyses that are robust against population stratification^7^. In particular, trio-based designs enable the analysis of transmission profiles, independent of the underlying genetic ancestry, given the parental genetic background. Specifically, these analyses compare the observed offspring genotypes with the expected offspring genotypes given the parental genetic background and Mendelian laws. This robust adjustment for parental genetic background is of particular interest in the context of PRS, since generalizability across populations is lacking and a major topic of current research^8^. Addressing this potential, Wang et al. recently introduced a methodological framework, PGS.TRI, to analyze PRS using case-parent trio designs^9^.

Another approach that aggregates genetic information to pinpoint potential causal factors in complex disease etiology is the integrative analysis of genetically predicted omics measurements, such as proteomics. Proteome-wide association studies (PWAS) have the potential to reveal new therapeutic targets^10^. The genetic prediction models underlying PWAS proteomics can be analyzed using case-parent trio designs, as in PRS analysis.

Here, we leveraged WGS data from two well-characterized family-based studies of childhood asthma: ‘Genetics of Asthma in Costa Rica Study’ (GACRS)^11^ and the ‘Childhood Asthma Management Program’ (CAMP)^12^, to perform trio-based PRS analyses using PGS.TRI and PWAS. We evaluated transmission patterns of polygenic scores derived from independent GWAS in the set of multi-ethnic trios, tested associations between the PRS and asthma-related clinical phenotypes, and conducted a two-step PWAS asthma analysis integrating external GWAS summary statistics.

Our analyses not only validate the utility of external asthma PRS in two independent family-based studies of childhood asthma but also provide new insights into their effects on clinical phenotypes and potential links between asthma and druggable proteins in the protein-based analysis.

## Results

### Study subjects for individual-level analyses

The individual-level data analyses utilized data from two family-based childhood asthma studies: GACRS and CAMP.

After genetic quality control, we retained 1,022 unrelated children with asthma from GACRS and 712 from CAMP. In addition, we identified 899 GACRS trios and 341 CAMP trios (all with affected children), of which 868 and 340 children, respectively, were also included in the unrelated-child analyses. Transmission testing of the polygenic risk scores and the individual-level data PWAS was performed using the trios; the clinical phenotype association analysis was performed using the larger set of unrelated children with asthma.

Table 1 reports and compares the demographic and clinical characteristics of unrelated children with asthma in GACRS and CAMP; the populations were similar with respect to sex, age, body mass index (BMI), and height. Significant differences are observed for self-reported race. All GACRS children are Hispanic, while most CAMP children are of European ancestry (around 10% Hispanic). We also observed significant differences in phenotype distributions between GACRS and CAMP, partly due to slightly different measurement targets; however, the measurement scales are comparable, enabling efficient meta-analysis of associations across these phenotypes. The clinical traits and phenotypes are further described and defined in Supplementary Table 1. Figures 1A and 1B visualize the highly comparable correlation structure between the phenotypes in GACRS and CAMP.

**Table 1.** Demographic and clinical characteristics of the cohorts of unrelated children with asthma in GACRS and CAMP.

|  | GACRS (n=1,022) | CAMP (n=712) | p-value |
| --- | --- | --- | --- |
| **Clinical demographics** |  |  |  |
| Sex (female) | 40% | 39% | p=0.64 |
| Age | 9.25 (1.86) | 9.07 (2.15) | p=0.07 |
| Body mass index | 18.30 (3.79) | 18.37 (3.48) | p=0.68 |
| Height(cm) | 132.80 (11.80) | 134.41 (13.81) | p=0.01 |
| Race | 100% Hispanic | 69% European, 13% African American, 10% Hispanic, 8% other | p<0.001 |
| **Parental history** |  |  |  |
| Maternal asthma | 31% | 27% | p=0.08 |
| Paternal asthma | 25% | 22% | p=0.10 |
| **Airway, atopy, and asthma severity** |  |  |  |
| L(P20) | 0.83 (0.58) | 0.05 (1.19) | p<0.001 |
| L(P20) binary | 16% | 5% | p<0.001 |
| Bronchodilator response binary | 14% | 27% | p<0.001 |
| Hay fever (yes or no) | 35% | 72% | p<0.001 |
| Hospital admission (yes or no) | 84% | 10% | p<0.001 |
| Log<sub>10</sub>(IgE) | 2.51 (0.66) | 2.65 (0.66) | p<0.001 |
| IgE (≥150) | 73% | 58% | p<0.001 |
| Log<sub>10</sub>(Eosinophil counts) | 2.61 (0.41) | 2.52 (0.52) | p<0.001 |
| Eosinophil counts (≥300) | 67% | 61% | p=0.009 |
| Skin prick test (positive numbers) | 3.07 (1.83) | 3.52 (2.61) | p<0.001 |
| Skin prick test (positive or negative) | 86% | 85% | p=0.56 |
| **Lung function** |  |  |  |
| FEV<sub>1</sub> (pre-BD) | 1.78 (0.50) | 1.66 (0.48) | p<0.001 |
| FEV<sub>1</sub> (post-BD) | 1.87 (0.51) | 1.83 (0.51) | p=0.11 |
| FVC (pre-BD) | 2.12 (0.59) | 2.11 (0.63) | p=0.79 |
| FVC (post-BD) | 2.16 (0.59) | 2.16 (0.64) | p=0.82 |
| FEV<sub>1</sub>/FVC (pre-BD) | 83.98 (7.92) | 79.33 (8.30) | p<0.001 |
| FEV<sub>1</sub>/FVC (post-BD) | 87.00 (6.86) | 85.23 (6.46) | p<0.001 |
| FEF<sub>2575</sub> (pre-BD) | 2.03 (0.74) | 1.57 (0.60) | p<0.001 |
| FEF<sub>2575</sub> (post-BD) | 2.34 (0.78) | 2.05 (0.65) | p<0.001 |
| FEF<sub>2575</sub>/FVC (pre-BD) | 97.24 (30.04) | 76.24 (26.65) | p<0.001 |
| FEF<sub>2575</sub>/FVC (post-BD) | 110.62 (30.65) | 97.69 (28.57) | p<0.001 |

GACRS: The Genetics Epidemiology of Asthma in Costa Rica Study; CAMP: Childhood Asthma Management Program. L(P20): Result of a methacholine challenge test (see Supplementary Table 1); BD: Bronchodilator; IgE: Immunoglobulin E; FEV<sub>1</sub>: Forced

### Transmission analysis of the external polygenic risk scores for asthma

Our analysis incorporated 7 asthma-related PRS, all of which were trained and constructed using independent external data from case-control studies. The first PRS was built as part of the multi-ancestry meta-analysis of asthma by the GBMI^5^, here denoted by GBMI-PRS. The second genome-wide PRS was derived by Moll et al.^14^ using the GWAS results from the TAGC^6^, in the following denoted by TAGC-PRS. Moreover, we investigated the five local PRS derived by Stikker et al. using epigenomic partitioning ^13^. Stikker et al. used H3K4me2 annotations across 19 cell types to partition asthma PRS variants into five regulatory clusters, defining cell-type-specific PRS components with distinct disease associations^2^. These PRS will be denoted by EGP-PRS-1 to EGP-PRS-5.

The PRS were computed for all trio members and for the set of unrelated children with asthma in GACRS and CAMP. The final number of genetic variants included in each PRS is reported in Supplementary Table 2. Figures 1C and D visualize the correlation structure between the seven PRS in GACRS and CAMP, showing the stronger correlation between GBMI-PRS and TAGC-PRS (as expected for genome-wide scores), as well as the weaker correlations with and between the EGP-PRS due to the partitioning.

Next, we tested the transmissions of these PRS using the PGS.TRI methodology proposed by Wang et al.^9^ (see Methods for more details). These analyses included testing for overall over-transmission, indirect parental effects, and interactions with parental asthma status and sex. Table 2 reports the corresponding results for the main effect analysis; Supplementary Table 3 contains all results, including indirect and interaction effects.

**Table 2.** Results of the transmission analysis of seven different polygenic risk scores in GACRS and CAMP. All polygenic risk scores were trained on external independent data. GACRS: The Genetics Epidemiology of Asthma in Costa Rica Study; CAMP: Childhood Asthma Management Program; GBMI: Global Biobank Meta-analysis Initiative; TAGC: Trans-National Asthma Genetic Consortium; EGP-PRS: polygenic risk score based on epigenomic partitioning as introduced by Stikker et al.^13^. The p-values are not adjusted for multiple testing.

| PRS | Effect<br>direction<br>GACRS | p-value<br>GACRS | Effect<br>direction<br>CAMP | p-value<br>CAMP | Effect<br>direction<br>GACRS+CAMP | p-value<br>GACRS+CAMP |
| --- | --- | --- | --- | --- | --- | --- |
| GBMI-PRS | + | 2.14e-16 | + | 1.34e-11 | + | 9.99e-25 |
| TAGC-PRS | + | 1.07e-09 | + | 1.15e-06 | + | 3.74e-14 |
| EGP-PRS-1 | + | 2.25e-02 | + | 1.84e-01 | + | 8.27e-03 |
| EGP-PRS-2 | + | 4.26e-02 | + | 8.56e-04 | + | 1.23e-04 |
| EGP-PRS-3 | + | 3.80e-12 | + | 6.39e-02 | + | 2.98e-12 |
| EGP-PRS-4 | + | 5.87e-01 | + | 2.74e-01 | + | 3.51e-01 |
| EGP-PRS-5 | + | 3.39e-01 | + | 1.73e-02 | + | 3.74e-02 |

We observed highly significant over-transmissions for GBMI-PRS and TAGC-PRS in the combined GACRS+CAMP analysis (p=9.99e-25 and p=3.74e-14, respectively). Among the EGP-PRS, EGP-PRS-3 obtained the smallest p-value with p=2.98e-12, which contains the well-established 17q21 asthma risk locus. The results show consistent findings in both GACRS and CAMP. Parental indirect effects, sex-interaction, and parental asthma interaction did not yield significant results after multiple testing correction (Supplementary Table 3). The joint analysis of GACRS and CAMP trios also did not reveal significant differences between cohorts (GACRS vs. CAMP) and no significant interaction effects with self-reported race after multiple testing correction (Supplementary Table 4), although nominally significant results were observed.

### PRS associations with clinical phenotypes

The next component of our analysis investigated the associations between PRS and clinical phenotype/variables in the set of unrelated children with asthma. Given the highly significant over-transmission or, respectively, association p-value, we focused on the GBMI-PRS. The analyses were performed separately in GACRS and CAMP, followed by a meta-analysis for each phenotype. Figure 2 and Supplementary Table 5 report the results of these analyses. The meta-analysis of GACRS and CAMP showed that 18 of 21 (86%) asthma phenotypes had nominally significant associations with GBMI-PRS, all of which showed a consistent direction of effect in both studies. After correction for 21 tests using the Bonferroni method, 13 p-values (62%) remained significant. In particular, an increasing GBMI-PRS was associated with increased allergen markers, such as skin prick test positivity, in both studies (p=4.5e-04 in GACRS and p=0.047 in CAMP). The same applies to Immunoglobulin E (IgE) levels, bronchodilator response, and measurements of increased eosinophil count (p=2.1e-07, p=1.0e-05, and p=8.2e-06, respectively, in the GACRS+CAMP meta-analysis). This suggests that the PRS identifies individuals with biomarkers of disease activity and type 2 inflammation. We evaluated the association between increased GBMI-PRS and increased inhaled corticosteroid use in GACRS and observed a nominally significant association (p=0.04, one-sided test). We did not perform this analysis in CAMP because steroid treatment was randomized in the trial. On the other hand, an increasing GBMI-PRS was associated with a decrease in lung function across several different metrics (Figure 2 and Supplementary Table 5). Additionally, we repeated the analysis using the TAGC-PRS, since the GBMI-PRS and TAGC-PRS are correlated, but not highly correlated, and potentially capture distinct characteristics. The results of this analysis are reported in Supplementary Table 6. The findings are qualitatively similar but yielded less significant associations. Supplementary Table 6 also contains results from an analysis in which both GBMI-PRS and TAGC-PRS were jointly tested for association with the phenotypes (in GACRS and CAMP separately, as well as meta-analyzed), yielding qualitatively similar results.

**Figure 1.**
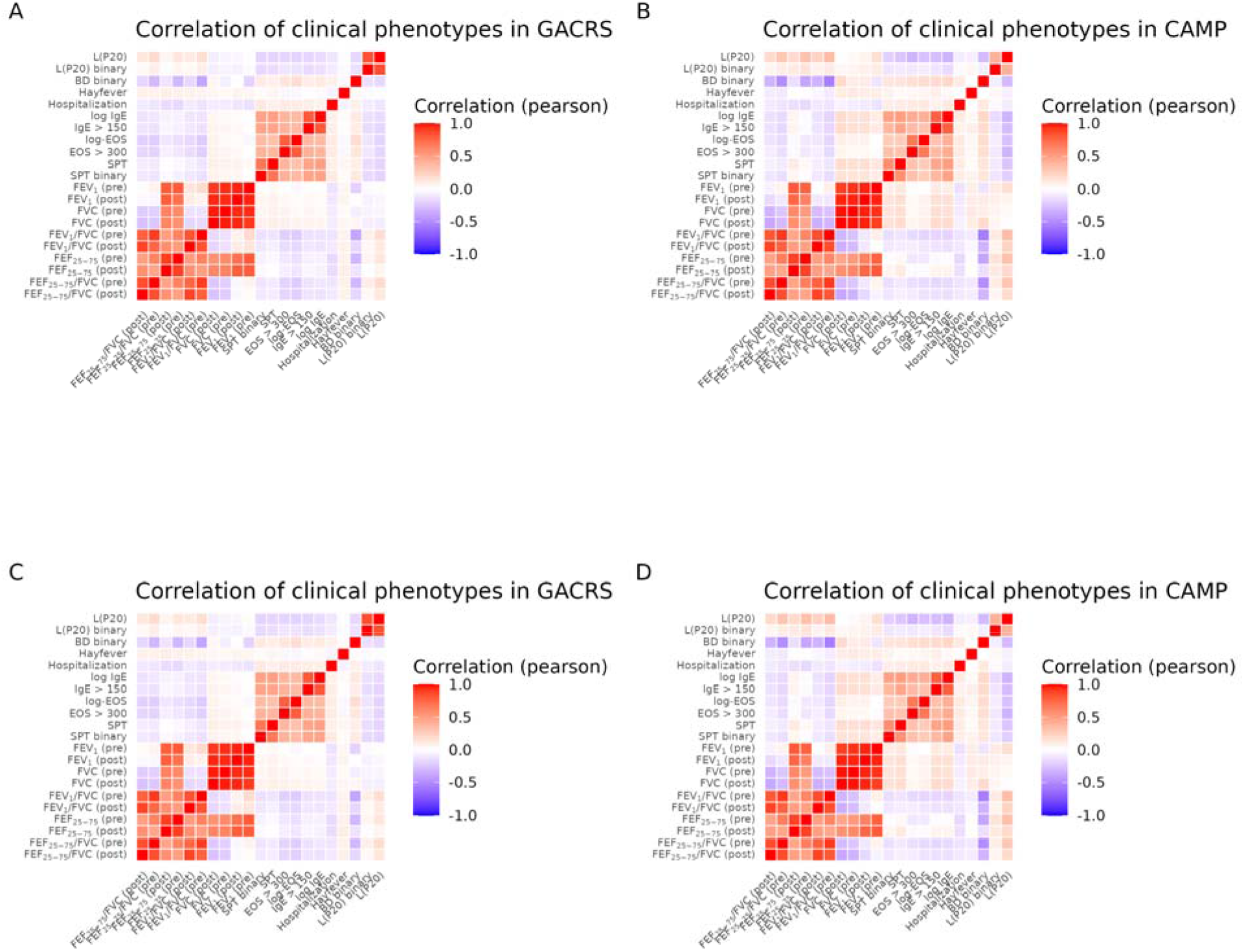
Correlation heatmaps for clinical phenotypes and polygenic risk scores in GACRS and CAMP. A and B: Correlation heatmap for the 21 clinical phenotypes and variables, calculated separately in GACRS (A) and CAMP (B). C and D: Correlation heatmap for the seven polygenic risk scores based on the set of unrelated children with asthma, separately in GACRS (C) and CAMP (D). PRS: polygenic risk score; GACRS: The Genetics Epidemiology of Asthma in Costa Rica Study; CAMP: Childhood Asthma Management Program. GBMI: Global Biobank Meta-analysis Initiative; TAGC: Trans-National Asthma Genetic Consortium; EGP-PRS: polygenic risk score based on epigenomic partitioning as introduced by Stikker et al.^13^.

**Figure 2.**
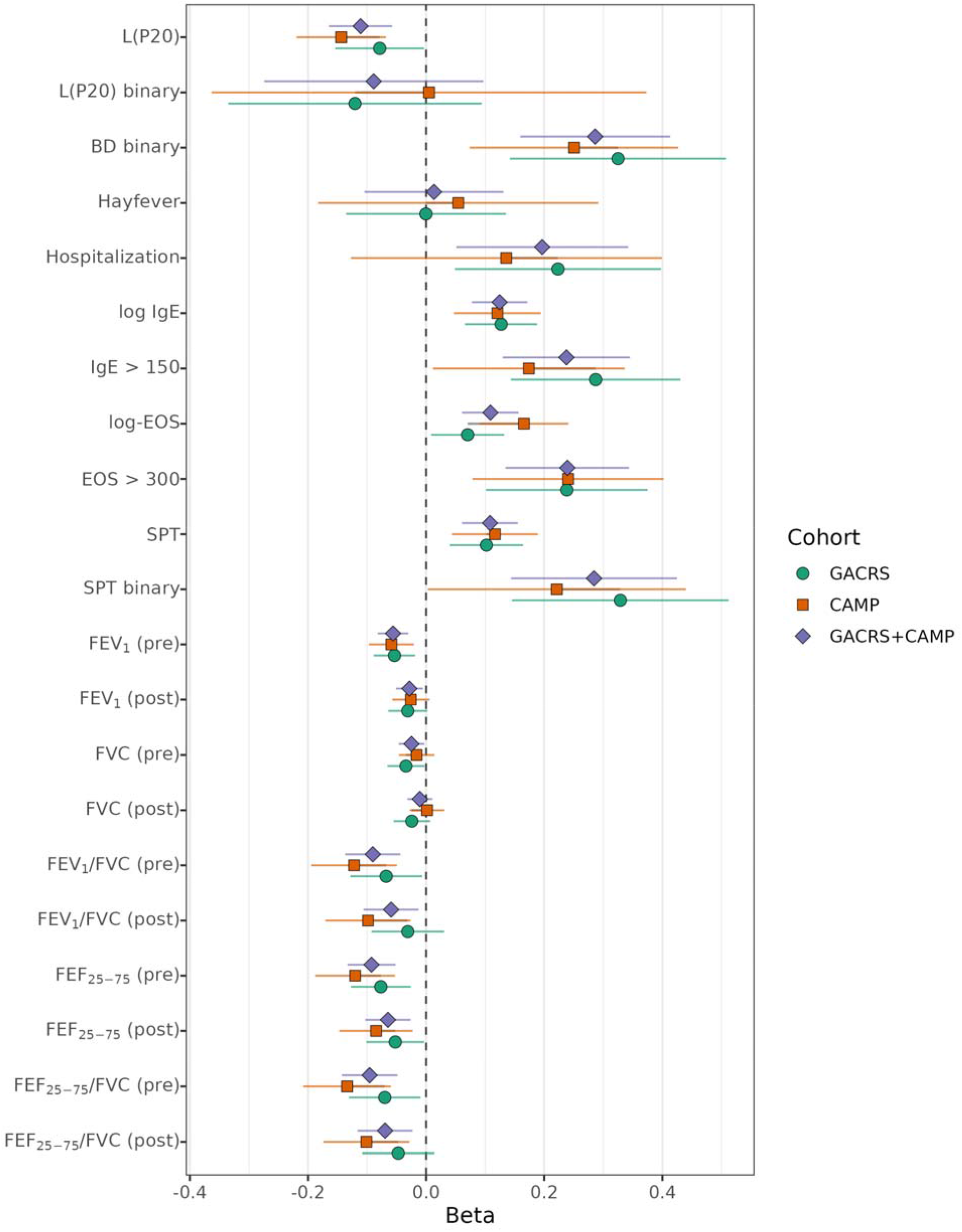
Effect size estimates corresponding to the association between the GBMI-PRS and the clinical phenotypes. The error bars correspond to the estimated 95% confidence intervals. Effect sizes are plotted for GACRS, CAMP, and GACRS + CAMP (meta-analysis). GACRS: The Genetics Epidemiology of Asthma in Costa Rica Study; CAMP: Childhood Asthma Management Program. L(P20): Result of a methacholine challenge test (see Supplementary Table 1); BD: Bronchodilator; IgE: Immunoglobulin E; SPT: Skin prick test; FEV_1_: Forced Expiratory Volume in 1 second; FVC: Forced Vital Capacity; FEF_2575_: Forced Expiratory Flow between 25% and 75% of Forced Vital Capacity.

### PWAS

The PRS analyses tested for over-transmission of the genome-wide and epigenetically partitioned genetic risk scores in case-parent trios. Following this idea, we investigated the transmission profiles of protein genetic scores. A significant under- or over-transmission in children with asthma indicates an association of the protein with asthma, essentially performing a proteome-wide association study (PWAS)^10^. Since the number of trios in the studies is limited and the statistical power is restricted, we implemented a two-step approach. The first step performs a summary-statistic-based PWAS using external data, and the second step tests the identified candidates on GACRS and CAMP data.

In particular, we included the genetic prediction models for 1,348 proteins, trained using plasma measurements and samples of European ancestry ^15^. Using the PWAS pipeline (see code and data availability), we obtained the PWAS p-values for 1,342 proteins based on the UK Biobank GWAS summary statistics for childhood-onset asthma (6 proteins were not sufficiently covered by the summary statistics)^2^. We considered the childhood-onset-asthma-specific GWAS results instead of the GBMI summary statistics to increase sensitivity. A total of 18 proteins were significant after Bonferroni correction for 1,342 tests at a significance level of 0.05. The results are consistent with findings in other recent studies that used similar GWAS summary statistics and the PWAS pipeline ^16,17^. For these 18 proteins, we evaluated the association p-value based on predicted protein levels using individual-level genetic data from GACRS and CAMP, employing the same approach as in the PRS transmission analysis. Additionally, we applied the PWAS pipeline approach to the association summary statistics obtained from a previous family-based genome-wide association study of asthma in GACRS and CAMP, using a largely overlapping set of trios^18^. The results are reported in Table 3 and Supplementary Table 7. We observed significant confirmation of the PWAS association of IL1R1 (p=0.045, corrected for 18 tests, consistent direction of effect) in the second step, in line with recent research highlighting this protein in the development of asthma ^16,17,19^. As the next step, we investigated the association between predicted IL1R1 protein levels and the 21 clinical traits and phenotypes.

**Table 3.** Results of the two-step PWAS using the UK Biobank asthma GWAS summary statistics (first step) and the second-step analysis in GACRS + CAMP. UKB: UK Biobank; GACRS: The Genetics Epidemiology of Asthma in Costa Rica Study; CAMP: Childhood Asthma Management Program. The p-values shown are not adjusted for multiple testing.

| Protein | CHR | NSNP | UKB PWAS Z | UKB PWAS p-value | GACRS+CAMP Z | GACRS+CAMP p-value |
| --- | --- | --- | --- | --- | --- | --- |
| IL6R | 1 | 49 | 4.90 | 9.50e-07 | 1.11 | 2.67e-01 |
| S100A7 | 1 | 37 | 4.70 | 2.63e-06 | -1.25 | 2.11e-01 |
| IL1R1 | 2 | 63 | 5.48 | 4.33e-08 | 3.02 | 2.50e-03 |
| IL1RL2 | 2 | 59 | -7.60 | 3.02e-14 | -1.45 | 1.48e-01 |
| IL1RL1 | 2 | 54 | -7.82 | 5.10e-15 | -0.75 | 4.51e-01 |
| TLR1 | 4 | 22 | -11.60 | 4.29e-31 | -1.22 | 2.22e-01 |
| IL7R | 5 | 16 | 4.28 | 1.84e-05 | 1.31 | 1.89e-01 |
| ATF6B | 6 | 86 | 7.13 | 9.70e-13 | -0.77 | 4.41e-01 |
| MICA | 6 | 158 | 6.97 | 3.09e-12 | 0.61 | 5.45e-01 |
| NCR3 | 6 | 107 | 5.84 | 5.09e-09 | -0.05 | 9.58e-01 |
| AGER | 6 | 17 | -10.48 | 1.08e-25 | -0.76 | 4.45e-01 |
| MICB | 6 | 159 | -5.93 | 3.04e-09 | 1.93 | 5.37e-02 |
| HLA-DQA2 | 6 | 83 | -10.24 | 1.34e-24 | 0.10 | 9.19e-01 |
| MRVI1 | 11 | 65 | 4.38 | 1.18e-05 | -1.50 | 1.34e-01 |
| ERBB3 | 12 | 60 | -4.26 | 2.05e-05 | -1.16 | 2.46e-01 |
| HEXIM1 | 17 | 29 | 6.30 | 2.94e-10 | -0.60 | 5.45e-01 |
| HMHA1 | 19 | 24 | 5.19 | 2.09e-07 | 1.83 | 6.75e-02 |
| KLK11 | 19 | 35 | -4.67 | 3.07e-06 | -1.20 | 2.30e-01 |

The results of this analysis are summarized in Supplementary Table 8. The meta-analysis between GACRS and CAMP did not yield any significant findings, but we observed nominally significant associations with lung function in GACRS.

Although IL1RL1, IL1RL2, and TLR1 did not reach statistical significance in the combined GACRS/CAMP analysis, these genes showed consistent effects in the same direction as the UK Biobank results. The three genes are functionally related to the IL-1 inflammatory signaling pathway: IL1RL1 (ST2) is the receptor for IL-33, a member of the IL-1 cytokine family ^20,21^, IL1RL2 forms part of the receptor complex for IL-36 cytokines ^22^, and TLR1 participates in innate immune signaling pathways that overlap with IL-1–mediated inflammatory responses ^23^.

Finally, we attempted to identify heterogeneity among children with asthma based on genetically predicted protein levels. We extracted the predicted offspring and the expected (based on parental genotypes) predicted protein levels for the 1,344 available proteins in the combined GACRS + CAMP trio dataset. This allowed us to compute the Mendelian residuals of predicted protein levels. Next, we analyzed the expected predicted protein levels and the Mendelian residuals using a singular value decomposition. As illustrated in Supplementary Figure 1, the first two components of the expected predicted levels separate self-reported race and cohort, while these clusters disappear for the Mendelian residuals. This observation reflects the ability of the parental adjustment to correct for the genetic background/ancestry of the trio. On the other hand, there appears to be no apparent clustering of the Mendelian residuals that could indicate biologically relevant asthma endotypes.

## Discussion

In this study, we analyzed external asthma PRS using WGS data in two well-characterized family-based studies of childhood asthma. Using a recently developed trio-based PRS transmission testing approach, we demonstrated significant over-transmission of the PRS derived from the GBMI and TAGC meta-analyses. These findings confirmed that the PRS capture substantial parts of the genetic architecture of asthma in these multi-ethnic children with asthma. The trio-based PRS approach is robust to population stratification, strengthening the results of the analysis. Furthermore, our analysis did not reveal significant differences in transmission patterns between GACRS and CAMP, despite the differences in genetic ancestries between these studies and the broader recent discussion about the transportability of PRS across populations.

Moreover, we observed associations between the GBMI-PRS and multiple asthma-related clinical phenotypes, including increased atopic dermatitis, elevated IgE levels, increased eosinophil counts, and bronchodilator responsiveness, as well as decreased lung function across several spirometry indices. These observations suggest that the genetic score is identifying individuals with biomarkers of disease activity and type 2 inflammation. Consequently, we tested for an association between increased GBMI-PRS and increased inhaled corticosteroid use in GACRS and observed a nominally significant association with consistent direction of effect. Overall, the associations showed largely consistent effects in both GACRS and CAMP, highlighting the potential of asthma PRSs to capture biologically relevant dimensions of asthma heterogeneity.

At the molecular level, we used genetically predicted plasma proteomics and childhood-onset asthma GWAS from the UK Biobank to identify 18 proteins whose predicted abundance is associated with childhood asthma and then evaluated these proteins in our trio-based GACRS and CAMP datasets. Among these, IL1R1 was the only protein whose association replicated in the family-based analysis, providing convergent genetic evidence that IL-1 signaling contributes to pediatric asthma. IL1R1 encodes the type 1 interleukin-1 receptor, the canonical signaling receptor for IL-1α and IL-1β, which is expressed on multiple lung structural (e.g., epithelial, fibroblast, smooth muscle) and immune cell types ^24^. Through MyD88–dependent signaling, IL1R1 activation induces inflammatory cytokine and chemokine production and has been linked to airway inflammation and remodeling processes, thereby connecting genetic susceptibility to inflammatory pathways implicated in asthma, including neutrophilic and potentially mixed inflammatory phenotypes^25,26^. Our result is in line with other relevant studies. Wang et al. applied a Mendelian randomization (MR) framework leveraging pQTLs from plasma and brain and showed that higher genetically predicted IL1R1 levels increase asthma risk^17^. Chen et al. extended this approach to 734 circulating and 154 cerebrospinal fluid proteins, again identifying a causal relationship between IL1R1 and asthma in large European biobank datasets^16^. Complementing these findings, Donoghue et al. integrated biobank-scale genetics with plasma proteomics in the UK Biobank Pharma Proteomics Project and identified a broad set of proteins with genetically supported roles in asthma risk and heterogeneity, including IL1R1^19^. Given that IL1R1 is directly targeted by clinically approved IL-1 pathway inhibitors such as anakinra, these convergent genetic findings further support the potential for therapeutic repurposing of IL-1– targeted agents in asthma^17^.

Despite these strengths, our study has several limitations. First, although GACRS and CAMP are among the largest family-based asthma cohorts with whole-genome sequencing, the overall sample size remains modest compared with large-scale biobank studies. This may have limited our power to detect subtler transmission effects, transmission differences, or interaction effects, particularly for the sub-PRS. This also applies to the PWAS, where only one of the 18 identified proteins is replicated. Second, the proteome prediction models used in PWAS were trained in European-ancestry samples, raising concerns about reduced predictive accuracy in non-European cohorts. Third, the PWAS is testing a limited portion of the proteome and is not as comprehensive as the PRS in terms of its coverage. Additionally, plasma proteomics might be a poor proxy of tissue-specific asthma-related proteomics. Finally, although we identified robust associations with clinical phenotypes, we cannot establish causal mechanisms, and further functional studies are needed to clarify how specific loci and pathways contribute to asthma pathophysiology. However, we speculate that children with higher asthma PRS - who, in our data, show more marked airflow obstruction, exacerbations, and T2-skewed biomarkers - might respond disproportionately well to therapies targeting alarmins (TSLP, IL-33) or IL1RL1/ST2, such as Tezepelumab^27^ or astegolimab^28^. Prospective trials incorporating genetic stratification and biomarker profiling will be required to test whether asthma PRS can enrich for responders and thereby improve the cost-effectiveness of these advanced biologics.

In conclusion, this work provides a comprehensive evaluation of external asthma PRS in family-based childhood asthma studies, integrating trio-based transmission testing, phenotype-wide association analyses, and proteome-wide approaches. By demonstrating over-transmission of polygenic risk, linking PRS to clinically relevant asthma endotypes, and identifying IL1R1 as a replicated protein-level association, our study advances the understanding of how inherited genetic risk contributes to asthma susceptibility and heterogeneity.

## Methods

### Study subjects

The genetic analyses are based on WGS data from the two childhood asthma studies, ‘The Genetic Epidemiology of Asthma in Costa Rica’ study (GACRS) and the Childhood Asthma Management Program (CAMP).

CAMP was a multicenter clinical trial designed to determine the long-term effects of three inhaled treatments for childhood asthma, as described previously^12^. CAMP focused on trios of asthmatic children and their parents and only included children with mild to moderate persistent asthma. Asthma was defined by the presence of symptoms or by the use of an inhaled bronchodilator at least twice weekly or the use of daily medication for asthma.

Study details and inclusion criteria for GACRS were described previously^11,29^. In short, GACRS recruited children between 6 and 14 years of age with physician-diagnosed asthma and at least two episodes of respiratory symptoms (wheezing, cough, or dyspnea) or a history of asthma attacks in the previous year^11^. Additional selection criteria included at least six great-grandparents born in the Central Valley of Costa Rica.

GACRS and CAMP applied comparable protocols for phenotyping of subjects.

### Consent

Written parental consent and the participating child’s assent were obtained for GACRS and CAMP. The GACRS protocol was approved by the Mass General Brigham Human Research Committee at Brigham and Women’s Hospital (Boston, MA; protocol No. 2000P001130) and the Hospital Nacional de Niños (San José, Costa Rica). CAMP protocols were approved by the Mass General Brigham Human Research Committee at Brigham and Women’s Hospital (Boston, MA; protocol No. 2011P000710).

### Whole-genome sequencing data

WGS data for GACRS and CAMP were generated as part of the National Heart, Lung, and Blood Institute (NHLBI) Trans-Omics for Precision Medicine (TOPMed) program^30^. Further details on sequencing and data quality control are described on the TOPMed website (https://topmed.nhlbi.nih.gov/). Our analyses are based on Freeze 10 data aligned to the GRCh38 reference and data with a minimum sequencing depth of 10 reads. We removed genetic duplicates, samples involved in pedigree discrepancies, and sex mismatches, identified using the kinship-based inference for GWAS (KING) tool^31^ and PLINK1.9/2^32^. We identified complete trios and the largest set of unrelated offspring in both studies. For each set of samples, we extracted the corresponding genetic variation data and filtered out variants with missing rates above 2%.

### Computation of PRS

We included two genome-wide polygenic risk scores for asthma in our analysis. The first was derived based on the multi-ancestry meta-analysis of asthma as part of the Global Biobank Meta-analysis Initiative (GBMI)^5^. The corresponding score information was obtained from the polygenic score catalog^33,34^ (PGS ID PGS001782). The second genome-wide PRS was constructed by Moll et al.^14^ using the genetic association summary statistics from the Trans-National Asthma Genetic Consortium (TAGC)^6^. Additionally, we investigated the five sub-PRS derived by Stikker et al.^13^. These sub-PRS with non-overlapping variants were constructed from the genome-wide childhood-onset asthma association data by Ferreira et al.^2^ and epigenomic data. Using information about the included genetic variants, effect alleles, and the corresponding weights, the PRSs were computed for each set of samples (trios, unrelated children) using the --score command in PLINK2 with mean imputation for missing genotype observations.

### Testing of PRS and sub-analyses

The PRS were tested using the methodology by Wang et al.^9^ as implemented in the R package PGS.TRI (see code and data availability). This test can be seen as a generalization of the single variant Transmission Disequilibrium Test (TDT)^35^ to a genetic score. We performed the test in GACRS and CAMP separately, as well as in the combined dataset. The *PGS.TRI* package enables testing both the main effect and interaction effects. For interaction effects, we considered sex, maternal asthma status, and paternal asthma status. Additionally, we also performed tests for indirect effects, corresponding to paternal effects not mediated by inherited genetic factors. Lastly, we investigated differential transmission between the two studies (GACRS vs. CAMP) and its modification by self-reported race using interaction testing in the combined set of GACRS and CAMP trios.

### Clinical variables

In addition to the trio-based PRS transmission testing, we also investigated the association between the PRS and clinical traits and phenotypes in the set of unrelated children with asthma in GACRS and CAMP (due to the larger sample sizes). We included 21 asthma-related clinical traits and phenotypes in our analysis that were available in both studies and had comparable definitions (Supplementary Table 1). These traits largely overlap with phenotypes utilized in a recent metabolomic study in GACRS and CAMP^36^.

### Calculation of principal components of genetic ancestry

The association analysis between PRS and clinical phenotypes in the set of unrelated offspring with asthma was adjusted for principal components of genetic ancestry (PCs). The PCs were computed based on the Genetic Relationship Matrix using PLINK2, which was based on a linkage disequilibrium-pruned set of genetic variants (window size of 500 kb, step size of 50 markers, and pairwise r^2^ < 0.1) in GACRS and CAMP separately.

### Association analysis between asthma PRS and clinical phenotypes

The association analysis between the asthma PRS and clinical phenotypes in a set of unrelated children with asthma was performed using linear regression for quantitative traits and logistic regression for binary traits. Quantitative traits were standardized before analysis (mean-centering, unit variance). All regression models were adjusted for age, sex, body mass index, height, and the first five principal components of genetic ancestry. The analyses were performed separately in GACRS and CAMP. The significance of p-values was confirmed by permutation-based testing. Next, we meta-analyzed the results from both cohorts using a fixed-effects approach.

### Proteome-wide association study (PWAS)

We utilized genetic prediction models of plasma proteins to perform a proteome-wide association analysis of asthma using a two-step approach. The genetic prediction models were trained on samples of European ancestry^15^. In the first step, we applied the PWAS pipeline corresponding to the plasma proteome analysis publication by Zhang et al.^15^ (see code and data availability) in combination with the UK Biobank^37^ childhood-onset asthma GWAS summary statistics published by Ferreira et al.^2^. This analysis tests the association between the predicted protein levels for each protein and childhood-onset asthma. We applied a multiple testing correction using the Bonferroni method and a significance level of 0.05. In the second step, the significant proteins were tested using the GACRS and CAMP trio data combined via the *PGS.TRI* approach. The results in the second step were corrected for multiple testing using the Bonferroni correction and the number of candidate proteins from step one. Additionally, we ran the PWAS pipeline approach with GACRS+CAMP asthma summary statistics from a recent publication^18^ for a comparison analysis.

## Data Availability

The weights for the GBMI PRS were downloaded from the PGS catalog (PGS ID PGS001782, https://www.pgscatalog.org/score/PGS001782/). The weights for the five sub-PRS derived by Stikker et al.13 were obtained from the corresponding Supplementary Materials. The PWAS genetic prediction models and respective pipeline scripts were obtained as described on http://nilanjanchatterjeelab.org/pwas/. The PGS.TRI R package is available at https://ziqiaow.github.io/PGS.TRI/.

## Code and data availability

The weights for the GBMI PRS were downloaded from the PGS catalog (PGS ID PGS001782, https://www.pgscatalog.org/score/PGS001782/). The weights for the five sub-PRS derived by Stikker et al.^13^ were obtained from the corresponding Supplementary Materials. The PWAS genetic prediction models and respective pipeline scripts were obtained as described on http://nilanjanchatterjeelab.org/pwas/. The *PGS.TRI* R package is available at https://ziqiaow.github.io/PGS.TRI/.

## Acknowledgments

Molecular data for the Trans-Omics for Precision Medicine (TOPMed) program was supported by the National Heart, Lung, and Blood Institute (NHLBI). Genome Sequencing for ‘NHLBI TOPMed: The Genetic Epidemiology of Asthma in Costa Rica’ (phs000988.v4.p1) was performed at Northwest Genomics Center (HHSN268201600032I, 3R37HL066289-13S1). Genome Sequencing for ‘NHLBI TOPMed: Childhood Asthma Management Program (CAMP)’ (phs001726.v1.p1) was performed at the Northwest Genomics Center (HHSN268201600032I). Core support including centralized genomic read mapping and genotype calling, along with variant quality metrics and filtering were provided by the TOPMed Informatics Research Center (3R01HL-117626-02S1; contract HHSN268201800002I). Core support including phenotype harmonization, data management, sample-identity QC and general program coordination were provided by the TOPMed Data Coordinating Center (R01HL-120393; U01HL-120393; contract HHSN268201800001I).

## Declaration of AI and AI-assisted technologies in the writing process

During the preparation of this work, the author(s) used ChatGPT⍰in order to⍰check and improve grammar, language refinement, and literature review. After using this tool/service, the author(s) reviewed and⍰edited the content as needed and take(s) full responsibility for the⍰content of the⍰publication.

