## Supplementary Figure for "Polygenic risk scores associate with asthma phenotypes and proteomic analyses implicate IL1R1 in two family-based studies"

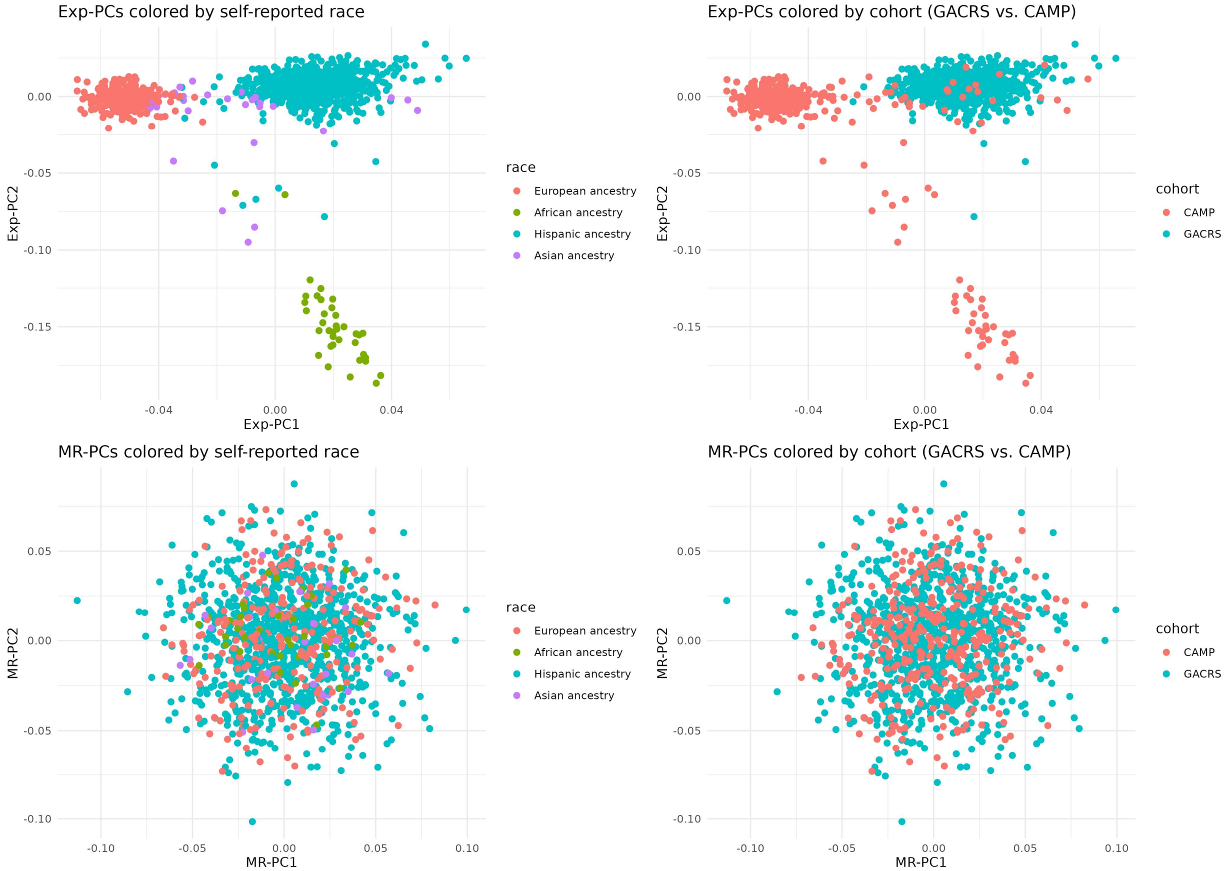


Supplementary Figure 1. Principal component plot for the 1,344 available predicted proteins. The upper row displays plots of the first two principal components of the matrix of expected protein levels, based on parental genetic data. The information is colored based on self-reported race (left) and study (right, GACRS vs. CAMP). The bottom row shows the corresponding plots based on the Mendelian residuals of predicted offspring protein levels adjusted for the expected levels (subtraction).
